# Evaluation Of Modified Hematological Sepsis Scoring System In Early Diagnosis Of Neonatal Sepsis

**DOI:** 10.1101/2025.02.02.25321553

**Authors:** Sishir Baruwal, Smriti Karki, Isha Gyawali, Smriti Khadka, Tek Nath Yogi, Santosh Maharjan

## Abstract

**Background:** Neonatal sepsis, a significant cause of morbidity and mortality, is a systemic response to infection in the first month of life. Its early diagnosis is challenging due to nonspecific clinical manifestations and the time-intensive nature of blood cultures, the diagnostic gold standard. The Hematological Sepsis Scoring (HSS) system, introduced in 1988, offers a rapid diagnostic alternative using hematological parameters. To enhance diagnostic accuracy, the Modified HSS incorporates additional parameters, such as nucleated RBC, and assigns higher weightage to neutropenia. This study evaluates the utility of the Modified HSS in the early diagnosis of neonatal sepsis, aiming to facilitate prompt treatment and reduce neonatal mortality.

**Methods:** This descriptive analytical study was conducted over a year at B.P. Koirala Institute of Health Sciences, involving neonates >34 weeks of gestation within four weeks of birth, clinically suspected of sepsis. Blood samples were analyzed using Modified Hematological Sepsis Scoring (HSS) and BacT/ALERT for culture. Purposive sampling selected 147 cases. Ethical clearance was obtained, and informed consent secured. Hematological parameters, including TLC, ANC, IT ratio, and nucleated RBC, were assessed, and cultures were processed for microbial identification and susceptibility.

**Results:** The study included 147 neonates with suspected sepsis (male:female ratio 1.33). Blood cultures were positive in 21 cases, with early-onset sepsis predominant (81%). Modified Hematological Sepsis Scoring (HSS) showed high diagnostic accuracy: sensitivity (90.48%), specificity (96.83%), and accuracy (95.91%) for HSS ≥4. Among hematological parameters, degenerative changes had the highest sensitivity (95.23%), and nucleated RBC showed the highest accuracy (91.15%). Staphylococcus aureus (52.3%) was the most common pathogen. Modified HSS proved effective for early sepsis diagnosis.

**Conclusion:** Modified HSS is an effective and accurate tool for early neonatal sepsis diagnosis, with high sensitivity, especially when using a score ≥4 as the cutoff. The inclusion of nucleated RBC enhances sensitivity, while the system’s diagnostic accuracy is improved by recalibrating parameters. Modified HSS, utilizing common hematological parameters, shows significant potential for clinical application.

## INTRODUCTION

Sepsis neonatorum is the term used to describe a systemic response to infection in newborn infants during the first month of life. The overall incidence of neonatal sepsis varies between 1% and 8% of all live births.[1] t includes systemic infections in newborns, such as meningitis, pneumonia, arthritis, and osteomyelitis. Bacteria may or may not be present during neonatal sepsis. Neonatal sepsis accounts for a significant proportion of early morbidity and mortality in neonates, with early-onset sepsis occurring within 24 hours of birth and late-onset sepsis occurring after 72 hours.[2]

Neonatal sepsis is more common in developing countries due to risk factors such as low birth weight, prematurity, low APGAR scores at birth, and poor socioeconomic conditions. Additional risk factors include premature rupture of membranes (PROM), chorioamnionitis or endometritis, Group B Streptococcal colonization, foul-smelling liquor, and meconium-stained amniotic fluid.[3] It is the most common cause of mortality, accounting for 22,500 deaths globally every year. Of the total sepsis-related neonatal deaths in 2013, 38.9% occurred in South Asia. The pooled incidence of culture-positive cases reported from South Asia is 15.8 per 1,000 live births. About 62% of infections in South Asia occur within the first 72 hours of life, roughly translating into an incidence of 9.8 per 1,000 live births.

In terms of pathogen profiles, there is a predominance of Gram-negative pathogens (>60%) and a low prevalence of Group B Streptococci in South Asia compared to the high incidence of Group B Streptococci in developed countries. Among isolates from hospital settings (n = 24,273), Gram-negative organisms (63%) were the most common, with Klebsiella spp. (23%), Escherichia coli (14%), and Acinetobacter (8%) being the top three. The most common Gram-positive organisms were Staphylococcus aureus (20%) and coagulase-negative Staphylococci (9%). [4] According to NDHS 2022, neonatal mortality rate of Nepal is 28 per 1,000 live births [5]. The incidence of neonatal sepsis in India is approximately 30 per 1,000 live births. The neonatal mortality rate in Bangladesh is 27 per 1,000 live births, with neonatal sepsis contributing to 36% of total neonatal deaths. The reported incidence of neonatal sepsis in Asia varies from 7 to 38 per 1,000 live births.[2]

The diagnosis of neonatal sepsis at its early stage is challenging, particularly due to its nonspecific clinical manifestations. Blood culture is considered the gold standard for diagnosing neonatal sepsis, with positive cultures obtained in 30–75% of cases. However, it requires 48–72 hours and a well-equipped laboratory.[6] To address the need for a rapid screening tool, R. L. Rodwell developed the Hematological Sepsis Scoring (HSS) system in 1988. This system utilizes various hematological parameters to enable early diagnosis and prompt initiation of treatment, thereby reducing neonatal morbidity and mortality. However, due to overlapping parameters, such as band count and immature-to-mature ratios (both representing the same pathological mechanism as the immature-to-total [I:T] ratio), a Modified Hematological Sepsis Scoring System (Modified HSS) was introduced. [7,8] The Modified HSS incorporates total leukocyte count (TLC), absolute neutrophil count (ANC), I:T ratio, degenerative changes, platelet count, and nucleated red blood cells (nRBC). It emphasizes the diagnostic significance of neutropenia over leukocytosis in neonatal sepsis, assigning a higher weightage (two points) to neutropenia. This adjustment enhances the diagnostic utility of the scoring system. The Modified HSS categorizes the likelihood of sepsis as follows: ≤2: Unlikely; 3–4: Probable; ≥5: Likely

The parameters used in the Modified HSS provide a practical framework for the early diagnosis of neonatal sepsis and have shown promise in improving clinical outcomes.[8] The different parameters used in Modified HSS for the diagnosis of Neonatal Sepsis can be explained as follows:

### Total Leucocyte Count (TLC)

Complete blood count (CBC) indices are most useful for detecting early-onset sepsis (EOS) after 4 hours. Variations in total leukocyte count are widely observed in sepsis. However, specific cutoff values for the normal range are not well-defined, limiting its diagnostic utility.[9]

### Absolute Neutrophil Count

Neutropenia is strongly associated with sepsis due to increased utilization of neutrophils at infection sites and their adhesion to endothelial cells. Recognizing its diagnostic significance, neutropenia was assigned a higher weightage (two points) in the Modified Hematological Sepsis Scoring System (Modified HSS).[8]

### Immature Cells

During sepsis, immature cells are released from the bone marrow, resulting in a "shift to the left." This leads to an increased proportion of immature-to-total (I:T) neutrophils. The I:T ratio is considered a more reliable predictor of neonatal sepsis compared to neutropenia or band count. However, in conditions like neonatal asphyxia, the I:T ratio may not rise, limiting its diagnostic utility in such cases.

### Platelet Count

A decline in platelet count is a common finding in sepsis. This decrease is likely attributable to disseminated intravascular coagulation (DIC) and the damaging effects of endotoxins.[10]

### Toxic Granules

Toxic granules are initial granules formed during the promyelocyte stage of neutrophil development. Composed of acidic mucosubstances and antimicrobial compounds, these granules enhance bactericidal activity by creating a more acidic environment in phagosomes, where bacteria are efficiently destroyed at a pH of 5.5 compared to a pH of 7.3. During inflammation, granulocyte colony-stimulating factor (G-CSF) levels rise, prompting neutrophils to produce toxic granules.[11]

### Degenerated forms

Neutrophilic vacuoles, often associated with toxic granules, are significant markers for sepsis. Similarly, Döhle bodies, comprising remnants of ribosomes and endoplasmic reticulum, are observed during bacterial infections. These appear as small, round or oval, light blue-grey inclusions in the peripheral cytoplasm of neutrophils. Döhle bodies are also seen in leukemoid reactions, inflammation, burns, after G-CSF administration, and during pregnancy.[11]

### Nucleated RBC

Nucleated red blood cells (nRBCs) are immature erythrocytes whose production is influenced by hypoxia, stress, and erythropoietin synthesis during sepsis. Sepsis is often characterized by an inappropriate immune response, with cytokine release driving increased nRBC production.[12] By incorporating nRBCs as a new parameter and assigning increased weightage to neutropenia, the Modified HSS has been applied for early diagnosis of neonatal sepsis. This approach has not been previously implemented in our setup, highlighting its novel utility.

## MATERIALS AND METHODS

### Type of Study Design

Descriptive Analytical

### Place of Study

Department of Pathology, Department of Microbiology and Department of Pediatrics.

### Duration of Study

1 year (Participants were prospectively recruited for this study between January 12, 2023, and January 12, 2024).

### Population/Participant’s selection criteria

i. **Inclusion Criteria:**

1. >34weeks of gestation
2. Baby within first four weeks of birth
3. Blood samples from clinically suspected cases of neonatal sepsis admitted in NICU in whom blood for culture and complete blood count had been sent in the same setting.
ii. **Exclusion Criteria:**

1. Cases unwilling to participate in the study
2. Cases in which blood culture was not sent
3. Cases with pathological jaundice

#### Study involved

Blood samples of clinically suspected cases of neonatal sepsis

#### Sample size was calculated using

Based on study conducted by Vani Krishnamurthy et al [8] 2018, sensitivity of Modified HSS was 0.84%

According to Buderer’s formula for total sample size:

N= Z2X Sensitivity(1-Sensitivity)/d2 X P

=1.96×1.96×0.84(1-0.84)/0.1×0.1×0.35

=3.84×0.1344/0.0035

=0.516096/0.0035

=147.45 i.e. 147

Where, Z = 1.96 in 95%CI

d = precision i.e. 0.10

#### Sampling Technique

Purposive sampling technique

#### Ethical Clearance

Ethical clearance was obtained from Institutional Ethical Review Board, (Ref no. 487/079/080-IRC, code number IRC/2319/022). Written informed consent was obtained from the parents or legal guardians of all neonatal participants before their inclusion in the study. The consent process included a detailed explanation of the study’s purpose, procedures, potential risks, and benefits. For illiterate guardians, the consent form was read aloud in the presence of an independent witness, who confirmed their voluntary participation with a signature. The study was approved by the Institutional Review Committee (IRC) and all procedures adhered to ethical guidelines.

#### Procedure

- Relevant history under the criteria for selection of sample was taken and recorded as per the proforma.
- After taking all aseptic precautions, 2ml of blood was withdrawn from all clinically suspected cases of neonatal sepsis who fulfilled inclusion criteria.
- 1ml of sample was sent for blood culture and sensitivity and another 1ml of the sample was anticoagulated with EDTA for Hematological study.

#### Procedure for Blood culture and sensitivity

One ml of blood was immediately inoculated to Blood culture bottles (Bact/Alert Paediatric aerobic bottles)

- BacT/ALERT 3D system:’

#### Principle

- Bact/ALERT bottles contain Tryptic soy broth and brain heart infusion broth added with adsorbent polymeric beads which neutralize the antimicrobials present in blood specimen.
- A sensor(liquid emulsion sensor) was bonded to the bottom of each bottle and separated from the broth medium by a differentially permeable membrane.
- CO2 produced by growing microorganisms diffuses across the membrane into the sensor where it reacts with water generating hydrogen ions.
- As the concentration of hydrogen ions increases and pH decreases, the blue green sensor becomes yellow, a change that was detected by colorimetric method.
- The algorithm for detection of growth is based on an analysis of the rate of change of CO2 concentration occurring in each bottle.
- Bottle which gives alert sound was taken for subculturing onto the blood agar and Mac Conky agar plates, isolation and identification of the growth and determination of antimicrobial susceptibility testing to commonly used antimicrobials in the institution was done by standard microbiological technique.
- Bottles which donot give beep sound was reported as culture negative at the end of 5 days.

#### Procedure for hematological study

- Values of total leucocyte count (TLC) and platelets count was noted using BeneSphere 5-part hematology analyser H51and counter checked manually.
- Peripheral smears was made and stained by Jenner-Giemsa stain and examined.

### Staining technique for Standard Giemsa Stain

- Smear was fixed in methanol for 5-7 min, then allowed to dry.
- Giemsa Stain was diluted 1:20 with deionized water
- Films were stained for 15-60min
- Rinsed in deionized water
- Air dryed and observed under microscope.
- Differential leucocyte count, Absolute neutrophil count, immature to mature neutrophil ratio and nucleated RBC was examined on stained smears.
- Modified Hematological Sepsis Scoring System as mentioned in table 1 used to calculate the score of each blood samples.
- IT ratio is calculated dividing the total immature count by total
- neutrophil count (including both mature and immature neutrophil count).
- Nucleated RBC was calculated by keeping the cut off value 5%.

**Table 1:** Modified HSS [8].

| **Parameter** | **Abnormality** | **Score** |
| --- | --- | --- |
| Total leucocyte count | <5000 | 2 |
|  | >25000(at birth) | 1 |
|  | >30000(12-24hrs) | 1 |
|  | >21000(day 2 onwards) | 1 |
|  | Normal | 0 |
| Total neutrophil count | No neutrophils | 2 |
|  | Increased/ decreased | 1/2 |
|  | Normal | 0 |
| Immature total neutrophil ratio(IT ratio) | >0.2 | 1 |
|  | <0.2 | 0 |
| Degenerative changes | Present(> or equal to 3) | 1 |
|  | Absent | 0 |
| Platelet count | >150000/cumm | 1 |
|  | <150000/cumm | 0 |
| Nucleated RBC | >5% | 1 |
|  | <5% | 0 |

IT, Immature to Total; RBC, Red Blood Cell

### Degenerative changes in neutrophils were graded as follows

**Table 2:** Scoring for Dohle Bodies or Vacoulization [13].

| Score | Neutrophils involved |
| --- | --- |
| 0 | None |
| 1+ | <25% |
| 2+ | 25-50% |
| 3+ | 51-75% |
| 4+ | >75% |

**Table 3:** Scoring for Toxic Granules.

| Scoring | Interpretation |
| --- | --- |
| 0 | Normal granulation |
| 1+ | Slight |
| 2+ | Dark granulations in 50% of neutrophils |
| 3+ | Very heavy granulations in most cells |
| 4+ | 'Gross' toxic granules obscuring nucleous |

## RESULTS

This study involved 147 clinically suspected cases of neonatal sepsis admitted at NICU. Of these 147, 84 (57%) were male and 63(47%) were female in the study. The male: female ratio, thus was 1.33: Blood culture of 21 cases yielded the growth of bacteria. 126 cases with negative blood culture results were selected as controls.

There were 17 (81%) neonates in the culture positive group who presented with early onset sepsis (age ≤3 days) and 4 (19%) presented with late onset sepsis (age >3days). In the culture negative group, 108 (85%) neonates were of age ≤ 3 days and 19 (15%) were more than 3 days old. Clinical Manifestations of Neonates as shown in the figure 1.

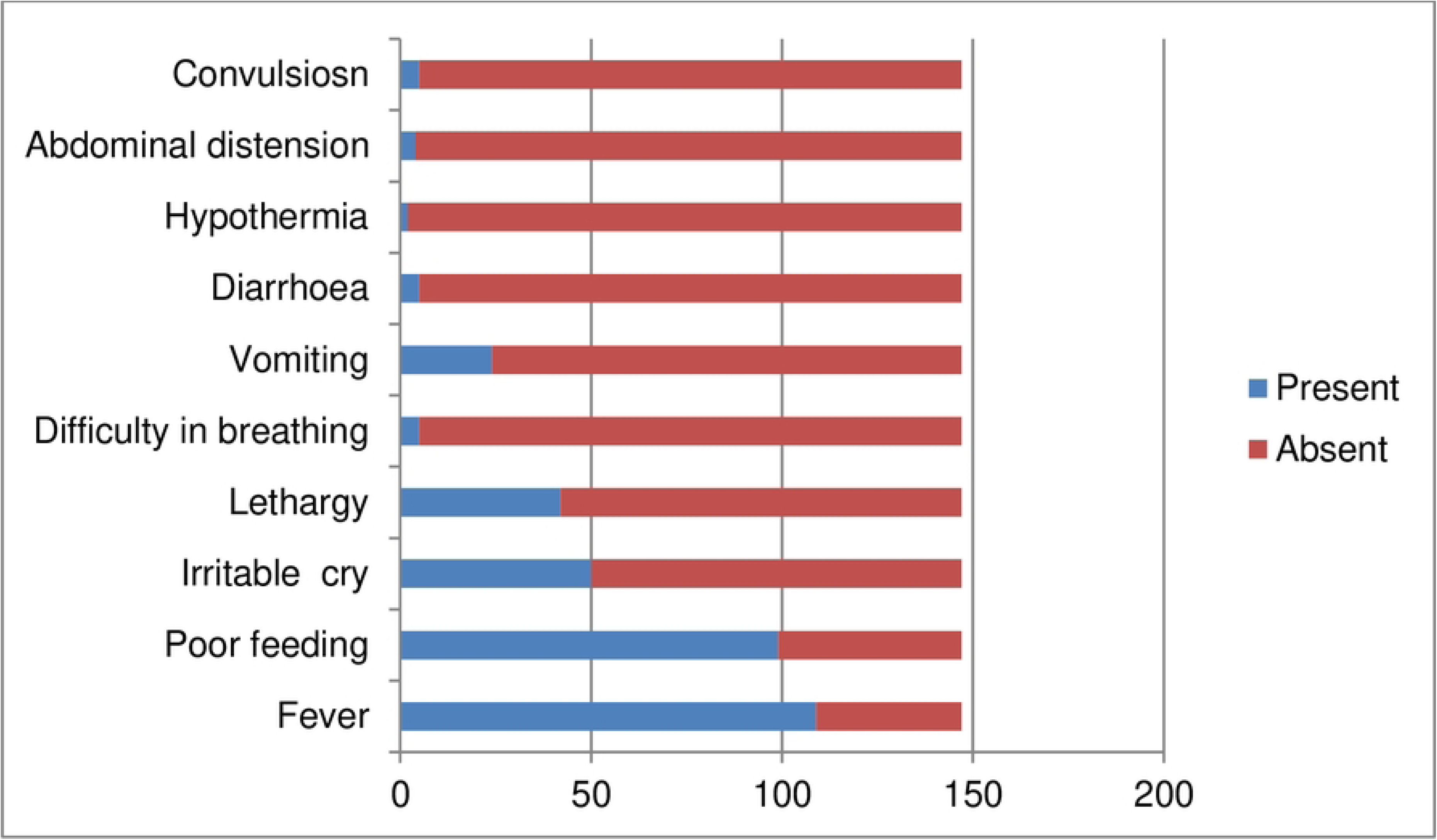

Neonates were mostly admitted with clinical manifestations of fever, poor feeding, lethargy 42(28%), irritable cry 50(34%), difficulty in breathing 5(3%), diarrhea 5(3%), convulsion 8(5.44%), abdominal distension 4(3%), hypothermia 2(1%), persistent vomiting 24(16%). Fever and poor feeding were present in majority of cases, i.e. 74(109)% and 67(99)%,respectively.

Table 4, 5: Out of the 147 cases, 16 were in the "sepsis likely" group, of which 14 (81%) were culture-positive; 37 were in the "probable sepsis" group, with 6 (19%) culture-positive; and 94 were in the "no sepsis" group, where only 1 (1%) case was culture-positive. Most culture-positive cases (11) scored ≥5 on the hematological scoring system (HSS), while the highest frequency of cases (34) scored 2.

**Table 4:**
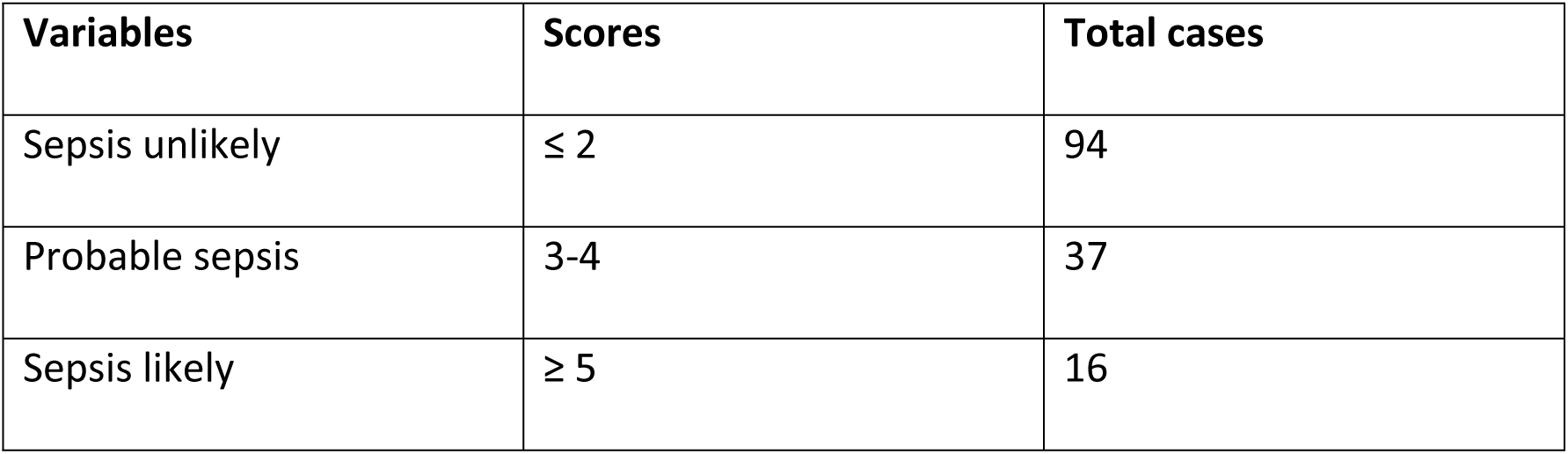
Modified Hematological Scoring System Category.

**Table 5:** Comparison of Modified HSS with culture results.

| Score | 0 | 1 | 2 | 3 | 4 | 5 | 6 | Total |
| --- | --- | --- | --- | --- | --- | --- | --- | --- |
| Culture positive | 0 | 0 | 1 | 1 | 5 | 11 | 3 | 21 |
| Culture negative | 32 | 28 | 33 | 29 | 2 | 2 | 0 | 126 |
| Total | 32 | 28 | 34 | 30 | 7 | 13 | 3 | 147 |

Table: 6: In culture-positive cases, 20 (95%) had HSS ≥3 compared to 23 (18%) in the culture-negative group. This yielded a sensitivity of 81.74%, specificity of 95.23%, positive predictive value (PPV) of 99.03%, negative predictive value (NPV) of 46.51%, and diagnostic accuracy of 83.67%.

**Table 6:** Strength of Modified HSS using Score ≥ 3 as cut off.

| Score | Culture negative | Culture positive |
| --- | --- | --- |
| <3 | 103 | 1 |
| $\geq 3$ | 23 | 20 |
| Total | 126 | 21 |

Table 7: With HSS ≥4, 19 (90%) culture-positive cases were identified, yielding a sensitivity of 90.48%, specificity of 96.83%, PPV of 82.60%, NPV of 98.39%, and diagnostic accuracy of 95.91%.

**Table 7:**
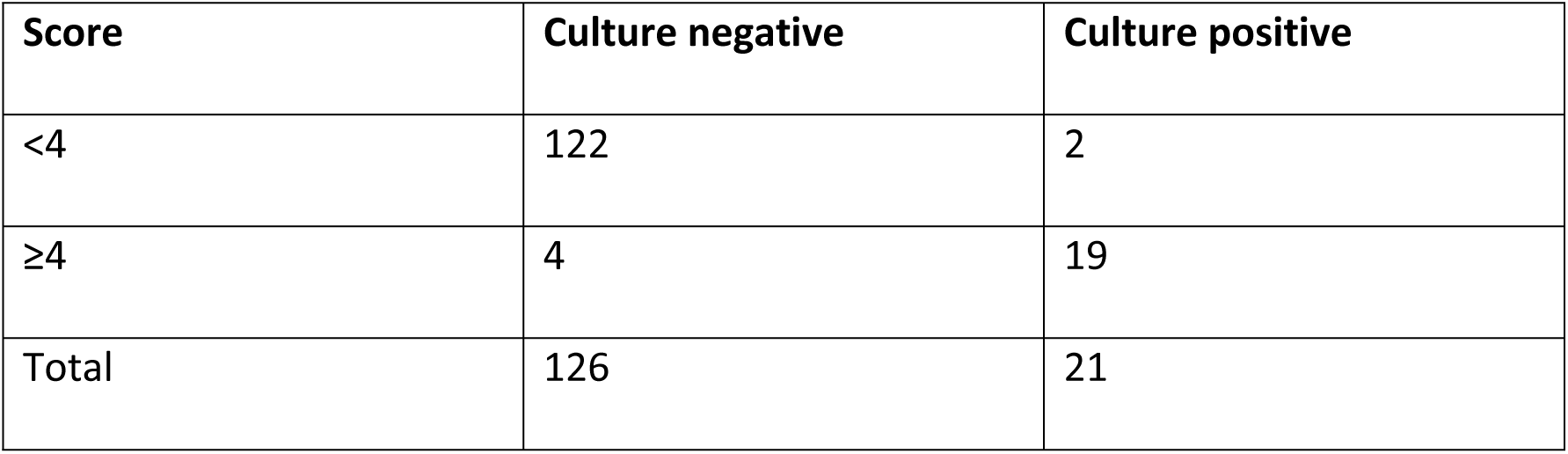
Strength of Modified HSS using Score ≥ 4 as cut off:

Table 8: Hematological parameters were assessed in both culture-positive and culture-negative groups. Values exceeding the normal range, contributing a score of 1, were classified as positive, while those within the normal range, contributing a score of 0, were classified as negative. Total leukocyte count (TLC) was positive in 10 cases (6.8% of the total), of which 6 were culture-positive. Total polymorphonuclear (PMN) count was positive in 90 cases (61.2%), with 19 culture-positive cases. The immature-to-total neutrophil (I:T) ratio was positive in 31 cases (21.1%), of which 13 were culture-positive. Degenerative changes were noted in 84 cases (57.1%), with 19 being culture-positive. Platelet counts were positive in 47 cases (32.0%), of which 15 were culture-positive. Nucleated RBCs were positive in 32 cases (21.8%), including 18 culture-positive cases.

**Table 8:** Individual hematological parameters in culture positive and culture negative group.

| **Hematological parameter** | **Cases with positive hematological parameter in Culture positive(n)** | **Cases with positive hematological parameter in Culture negative group (n)** | **Total cases with positive hematological parameters (n)** |
| --- | --- | --- | --- |
| TLC | 6 | 4 | 10 |
| ANC | 19 | 71 | 90 |
| IT Ratio | 13 | 18 | 31 |
| Degenerated changes | 19 | 65 | 84 |
| Platelet count | 15 | 32 | 47 |
| Nucleated RBC | 18 | 14 | 32 |

TLC, Total leukocyte count; ANC, Absolute Neutrophil Count; IT, Immature to Total.

Table: 9: The sensitivity, specificity, positive predictive value (PPV), negative predictive value (NPV), and accuracy of individual hematological parameters were calculated using the Pearson Chi-square test to determine statistical significance. Among the parameters, degenerative changes exhibited the highest sensitivity (95.2%), followed by ANC and nucleated RBC counts (90.5%). Total WBC count demonstrated the highest specificity (96.8%), followed by nucleated RBC count (91.3%). PPV was highest for total WBC count (63.6%), while NPV was highest for degenerative changes (98.6%), followed by nucleated RBCs (98.3%). All parameters were statistically significant (p < 0.01), except for platelet count (p = 0.04). The highest diagnostic accuracy was observed with nucleated RBC count (91.2%), while ANC had the lowest accuracy (50.3%). When these parameters were combined in the modified Hematological Scoring System (HSS), the diagnostic accuracy was 83.7% for a cutoff score of ≥3, increasing to 95.9% for a cutoff score of ≥4.

Among the pathogens identified, Staphylococcus aureus was the most common (11 cases, 52.3%), followed by Enterococcus spp. (4 cases, 19.0%), and Escherichia coli, Acinetobacter spp., and Klebsiella spp., each accounting for 2 cases (9.5%) as shown in blood culture isolate in figure 2.

**Table 9:**
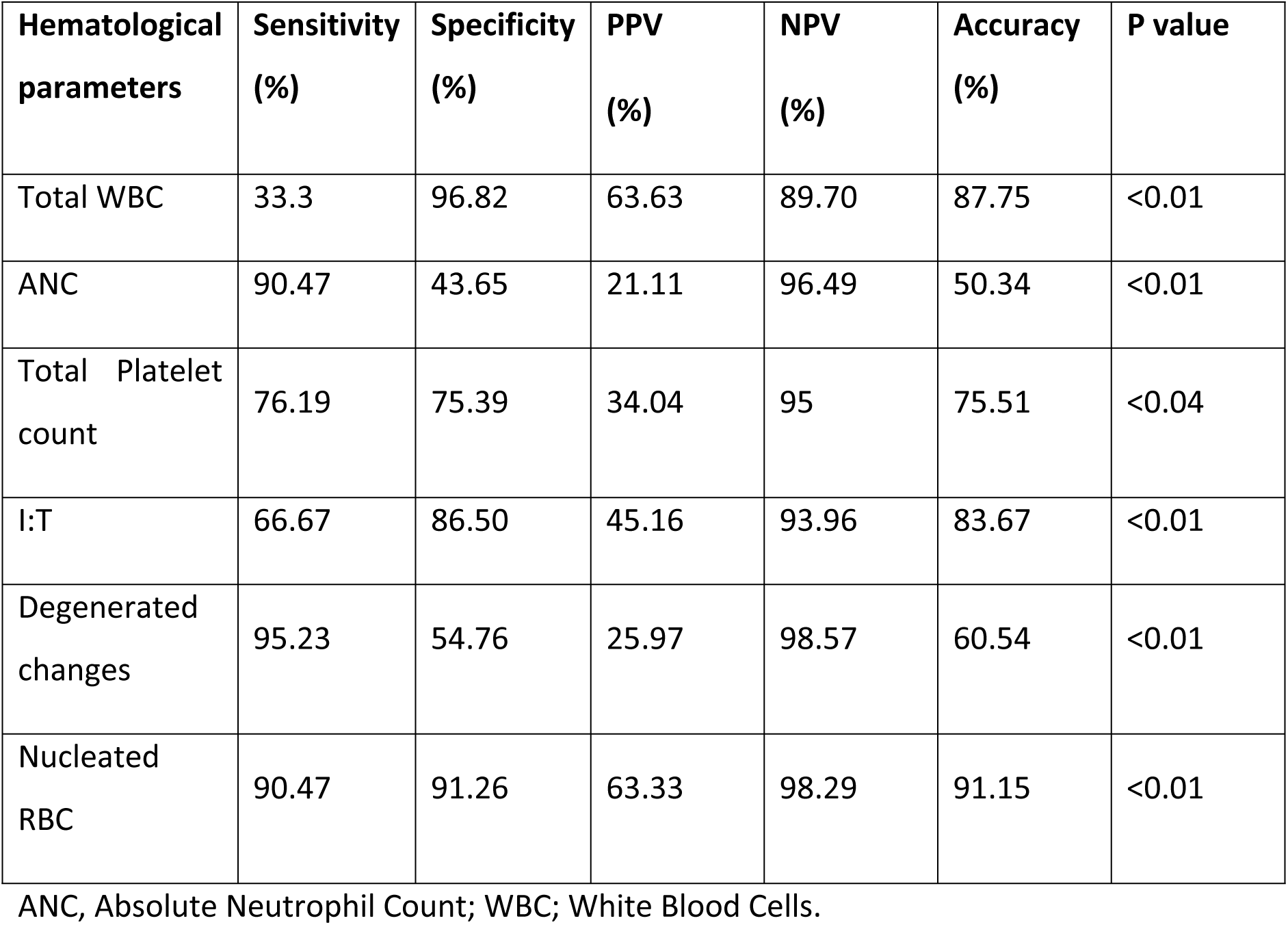
Strength of individual hematological parameters.

| **Hematological parameters** | **Sensitivity (%)** | **Specificity (%)** | **PPV (%)** | **NPV (%)** | **Accuracy (%)** | **P value** |
| --- | --- | --- | --- | --- | --- | --- |
| Total WBC | 33.3 | 96.82 | 63.63 | 89.70 | 87.75 | <0.01 |
| ANC | 90.47 | 43.65 | 21.11 | 96.49 | 50.34 | <0.01 |
| Total Platelet count | 76.19 | 75.39 | 34.04 | 95 | 75.51 | <0.04 |
| I:T | 66.67 | 86.50 | 45.16 | 93.96 | 83.67 | <0.01 |
| Degenerated changes | 95.23 | 54.76 | 25.97 | 98.57 | 60.54 | <0.01 |
| Nucleated RBC | 90.47 | 91.26 | 63.33 | 98.29 | 91.15 | <0.01 |

ANC, Absolute Neutrophil Count; WBC; White Blood Cells.

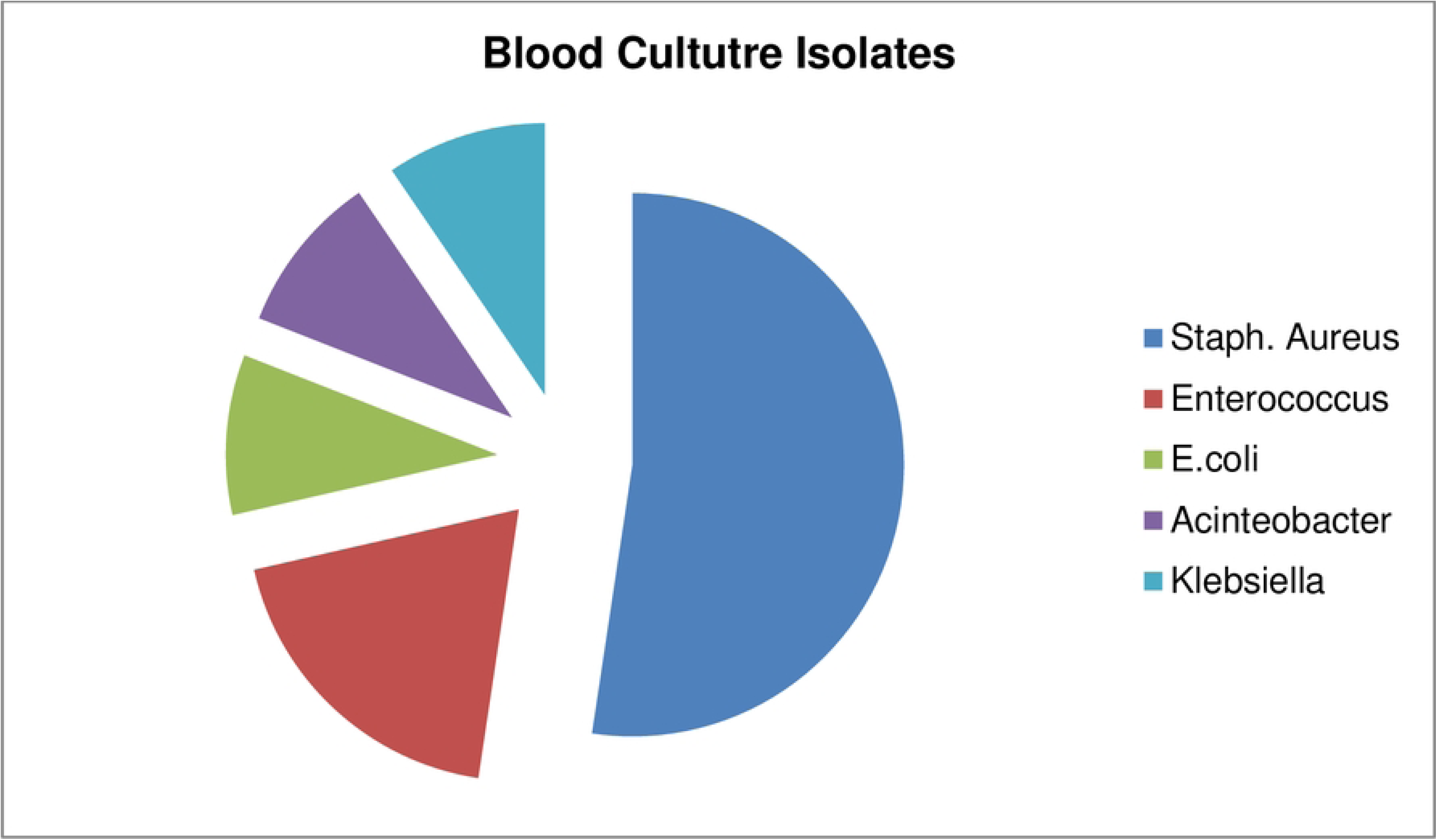

## DISCUSSION

This study aimed to facilitate the early diagnosis of neonatal sepsis by utilizing the Modified Hematological Scoring System (HSS) and comparing its diagnostic performance with Rodwell’s Hematological Scoring System. A novel parameter, nucleated red blood cells (NRBCs), was incorporated, and the weightage for low neutrophil counts was increased to a score of 2.

Nucleated red blood cells (NRBCs) are immature erythrocytes, the production of which is driven by hypoxia and erythropoietin (EPO) synthesis. EPO, a glycoprotein hormone, is synthesized by the kidneys, liver, spleen, lungs, and bone marrow. In human fetuses, the liver is the primary site of EPO synthesis, transitioning to the kidney closer to term. Other sites of production include the placenta, endothelium, and neuronal cells. Prior studies suggest that EPO increases erythroid production and releases immature erythrocytes into circulation in response to hypoxia.[14]

In our study, we included only the neonates who were more than 34 weeks of gestation and clinically suspected sepsis cases admitted in NICU setup for which both sample for complete blood count and culture was sent in same setting.

This study included neonates born after 34 weeks of gestation who were clinically suspected of having sepsis and admitted to the NICU. Complete blood count and blood culture samples were collected simultaneously. Among the 147 neonates studied, the incidence of sepsis was higher in males (57.14%) compared to females (42.85%), consistent with prior studies.[15] [16][6]. Early-onset sepsis (EOS) was more prevalent (85%) than late-onset sepsis (LOS) (15%), similar to findings from previous studies in Nepal.[2] [17][18]. However, Rasul et al. [18] reported differing definitions of EOS, ranging from 2 to 7 days, and noted an EOS incidence of 70.7% when defined as sepsis occurring within 7 days.

The clinical presentation of neonatal sepsis in this study was nonspecific, with fever and poor feeding being the most common complaints. Other symptoms included lethargy, irritability, difficulty breathing, diarrhea, convulsions, abdominal distension, hypothermia, or persistent vomiting. Blood culture remains the gold standard for confirmation, given the nonspecific nature of clinical symptoms.

Six hematological parameters were evaluated in this study, with a cutoff score of ≥4. Similar studies by Ahiraro et al.[19] (SN:95% SP:96%), Meirina et al [20] (SN :80% SP:90%)and Makkar et al. [21] (SN:100%, SP:74%) also used this cut off score where sensitivity and specificity was in higher range. Conversely studies by Adhikari et al [16] (SN:85.96%, SP :84.26%) Yusuf et al [3] (SN :71%, SP:72%) and Saleem et al [22](SN:90%,SP:74%) used the cut off value ≥3. In our study, out of 21 positive cases, 19 cases were above the cut off value ≥4, which showed that increase score more likely to be culture positive.

Among individual hematological parameters, NRBC emerged as the most reliable indicator, with a diagnostic accuracy of 91.15%, sensitivity of 90.47%, specificity of 91.26%, PPV of 63.33%, and NPV of 98.29%.Basner et al [12] suggested that hypoxia-driven EPO synthesis leads to increased NRBC production and that cytokine-mediated inflammatory responses further amplify NRBC release into circulation. Gwori et al.[23] did study to enumerate the role of nucleated in Neonatal Sepsis as an Early Response Element to Inflammation in Infection, where the sensitivity, specificity, PPV and NPV of nucleated RBC on identifying neonatal sepsis was 81.96%,61.11%,70.42% and 75% respectively. The mean value of NRBCs in culture positive sepsis was 13.69, in sepsis screen positive sepsis was 12.38 and 4.87 in no sepsis group of babies. Shalini et al.[15] Shalini et al. [15] (2023) compared HSS and a modified HSS that included NRBC (termed "sepscore") at a cutoff of ≥3, finding increased diagnostic performance with a 20% improvement in specificity.

Other parameters demonstrated variable diagnostic performance. Total leukocyte count (TLC) had low sensitivity (33.3%) but high specificity (96.82%), consistent with findings by Adhikari et al. [16] (SN: 38%, SP: 90%) and Vinay et al [24] and Elsayed et al [25] also found the sensitivity and specificity similar to our study as 49.6%, 60% and 58% respectively. Whereas the study done by Dey et al[26] in Bangladesh foun the TLC more sensitive with 97% and specificity of 47.85%.Khair et al pointed that TLC has been considered to be of less diagnostic value by the investigators of this study because of the wide variation in its value. However, they have also labeled it as good parameter for confirmation of sepsis.[27]

On peripheral smear examination,degenerative changes were seen as vacoules within the neutrophilic cytoplasm along with swelling of the nucleus with smudged chromatin. Degenerative changes had the highest sensitivity(95.23%) and NPV (98.57%) of all the parameters along with accuracy of only of 60.54%. However, the specificity was only 54.76% and PPV was 25.97%. Yusuf et al had pointed out that toxic granules are only seen during infection and stress induced leucopoiesis and are absent in healthy babies. However, the quantity of these granules may not always be increased even though their presence invariably indicates sepsis.[28]

In our study, platelet count had sensitivity, specificity, PPV and NPV of 76%, 75.39%, 45.16% and 93.96% respectively which was supported by study done by Derbala et al which showed NPV of 100%. Whereas study done Adhikari et al showed sensitivity, specificity, PPV and NPV 68%, 39.5%, 55.76% and 53.11% respectively. Decreased platelet count is seen in sepsis possibly because of increased destruction, sequestration due to infection or due to reduced production owing to the damaging effects on the megakaryocytes by the endotoxins.[10][25]

In our study, ANC showed sensitivity of 90.47 and NPV of 96.49%. However, the specificity and PPV are low, i.e. 43.65% and 21.11% respectively. Similarly, I: T ratio showed specificity (86.50%) and NPV (93.96%) but low sensitivity (66.67%) and NPV (45.16%). Band cells are the direct precursors of mature neutrophils. Mature neutrophils are responsible for the elimination of foreign bacteria.In sepsis, increase number of immature neutrophils reflect disease severity and patient deterioration.[14]

The slight variation in the results of hematological parameters among different studies might be due to difference in blood sampling time, exact method of test applied, diagnostic criteria followed, severity of infection and age of newborns. Comparison has not been done with those studies who have studied only selected number of hematological parameters and have not calculated a hematological score using Rodwell‟s Hematological Scoring System.

Our study pointed out the fact that all the parameters were statistically significant. The chances of culture positivity was more in the sepsis likely group and culture was negative in no sepsis group. It proves that higher the sepsis scoring, more the baby are in risk of sepsis. In our study, sensitivity, specificity, accuracy of Nucleated RBC was found to be 90.47%, 91.26% and 91.15%, respectively. Several studies(Shalini et al [15] 2023, Gwori et al [23] 2018) pointed the fact about the release of nucleated RBC in peripheral smear.They are more commonly release during stress and sepsis.Appearance of normoblasts in peripheral circulation is a serious concern. Their presence in the peripheral blood indicates that the bone marrow barrier has been disrupted or that extramedullary hematopoiesis has been triggered[15].The specificity and sensitivity of our study was 96.83% and 90.48% respectively. So, with our study, it was conclusive that, newly included parameter, nucleated RBC was significant for the early diagnosis of neonatal sepsis.

**Table 10:**
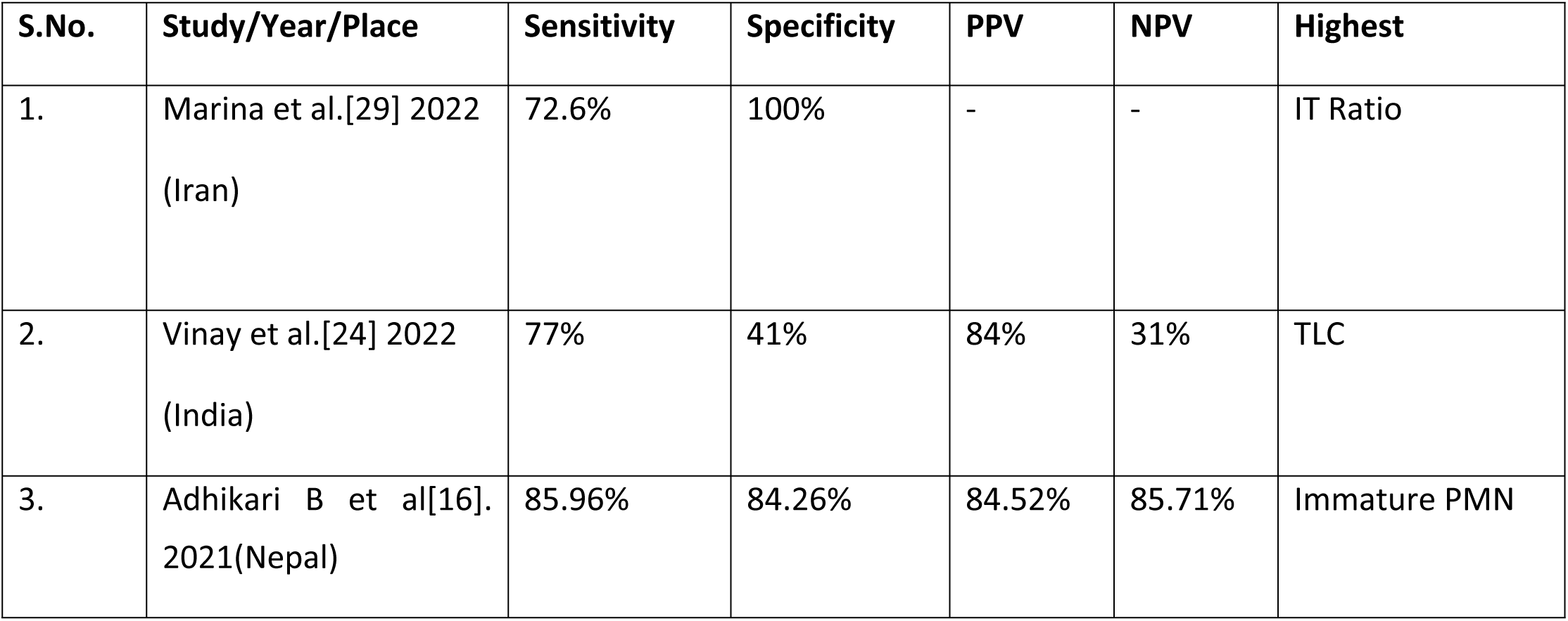
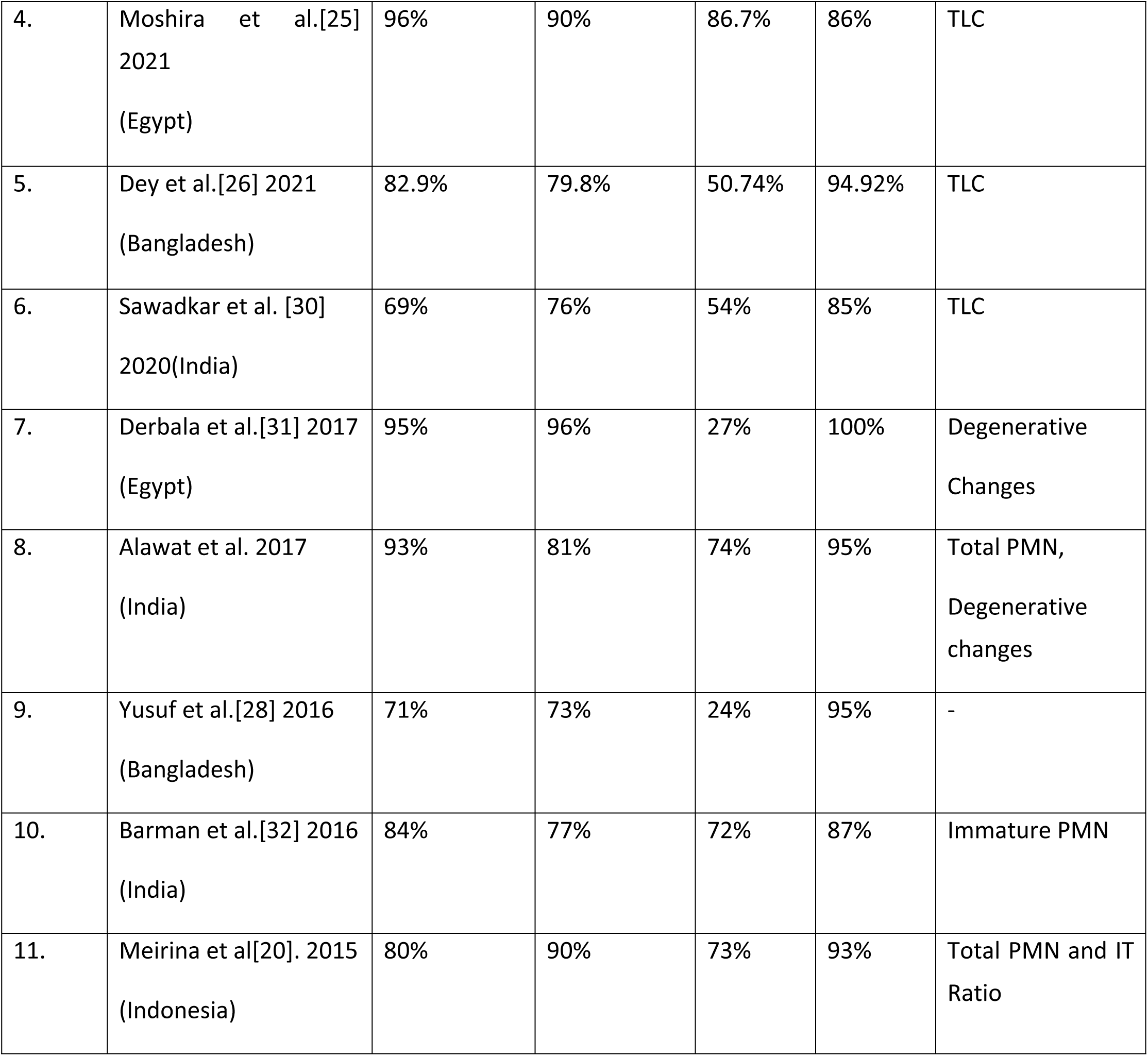
Comparison Between HSS of Different Studies.

| S.No. | Study/Year/Place | Sensitivity | Specificity | PPV | NPV | Highest |
| --- | --- | --- | --- | --- | --- | --- |
| 1. | Marina et al.[29] 2022<br>(Iran) | 72.6% | 100% | - | - | IT Ratio |
| 2. | Vinay et al.[24] 2022<br>(India) | 77% | 41% | 84% | 31% | TLC |
| 3. | Adhikari B et al[16].<br>2021(Nepal) | 85.96% | 84.26% | 84.52% | 85.71% | Immature PMN |
| 4. | Moshira et al.[25]<br>2021<br>(Egypt) | 96% | 90% | 86.7% | 86% | TLC |
| 5. | Dey et al.[26] 2021<br>(Bangladesh) | 82.9% | 79.8% | 50.74% | 94.92% | TLC |
| 6. | Sawadkar et al. [30]<br>2020(India) | 69% | 76% | 54% | 85% | TLC |
| 7. | Derbala et al.[31] 2017<br>(Egypt) | 95% | 96% | 27% | 100% | Degenerative<br>Changes |
| 8. | Alawat et al. 2017<br>(India) | 93% | 81% | 74% | 95% | Total PMN,<br>Degenerative<br>changes |
| 9. | Yusuf et al.[28] 2016<br>(Bangladesh) | 71% | 73% | 24% | 95% | - |
| 10. | Barman et al.[32] 2016<br>(India) | 84% | 77% | 72% | 87% | Immature PMN |
| 11. | Meirina et al[20]. 2015<br>(Indonesia) | 80% | 90% | 73% | 93% | Total PMN and IT<br>Ratio |

**Table 11:**
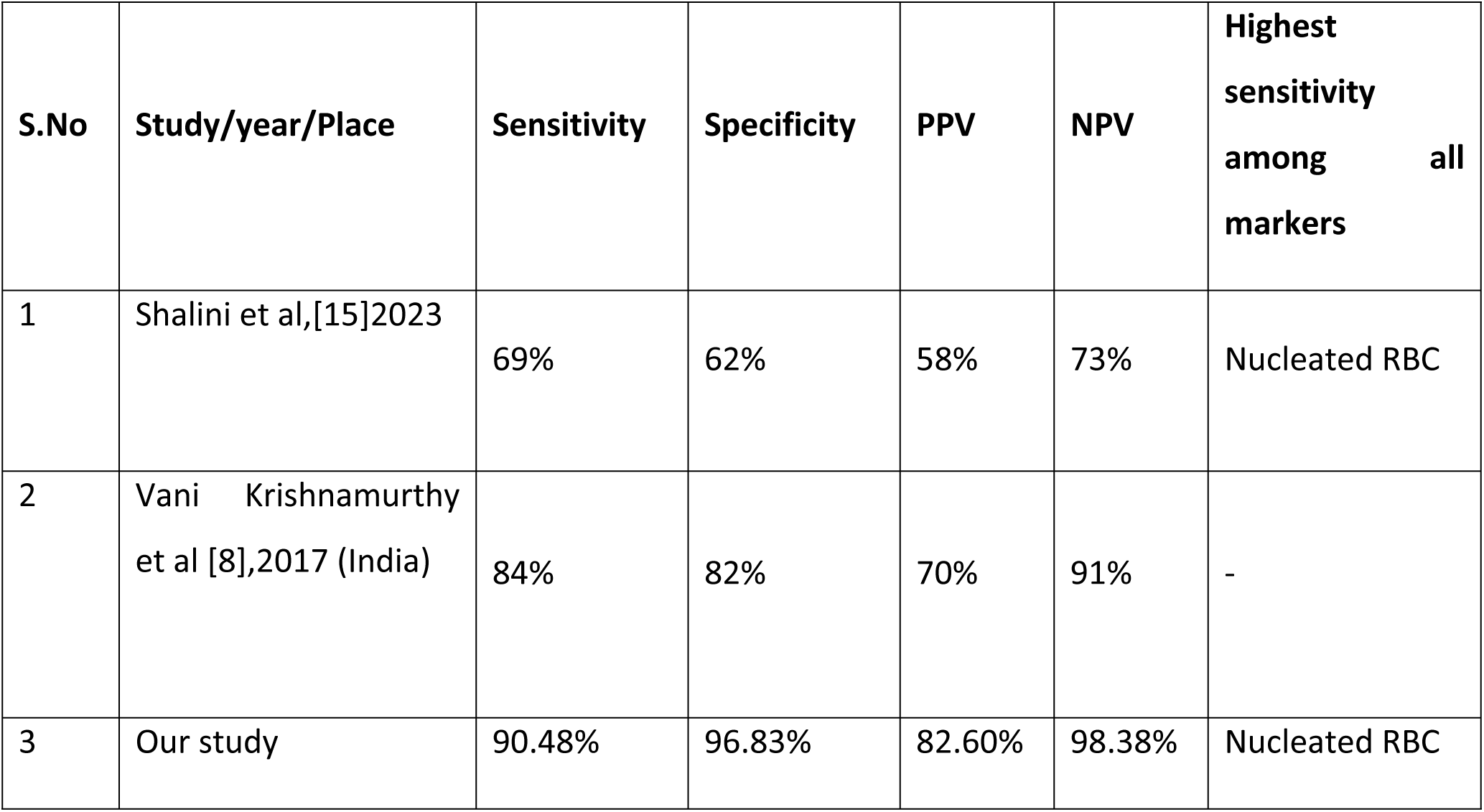
Various Modified HSS studies and our study.

| S.No | Study/year/Place | Sensitivity | Specificity | PPV | NPV | Highest sensitivity among all markers |
| --- | --- | --- | --- | --- | --- | --- |
| 1 | Shalini et al,[15]2023 | 69% | 62% | 58% | 73% | Nucleated RBC |
| 2 | Vani Krishnamurthy et al [8],2017 (India) | 84% | 82% | 70% | 91% | - |
| 3 | Our study | 90.48% | 96.83% | 82.60% | 98.38% | Nucleated RBC |

In our study, Staphylococcus Aureus was the most frequent (52.3%) organism to be isolated followed by Enteroccocus spp. (19.04%). This is similar but slightly different in comparison to the findings from the study by Adhikari et al [16] in Eastern Nepal where Acinetbacter spp was the next common organism to be isolated after Staphylococcus aureus. Study done by Khanal et al [33] 2004 showed the Gram positive isolates (73%) and Gram negative (27%), out of which Staphylococcus Aureus (60.2%) was most frequent organism to be isolated followed by Enterococcus(10.2%) which was similar to our study.

Similar study done by Yadav et al [34], also showed the Staphylocoocus Aureus (4.5%) as the common isolates in blood culture. A study done by Sanjana et al in Central Part of Nepal also showed that Staphylococcus Aureus remained the predominant isolate followed by Klebsiella spp. The incidence of gram positive and gram negative organisms were 44.1 % and 55.9 % respectively.

In study done by Lakhe et al [2] 20 also, Staphylococcus aureus(41%) was the most common isolate.But other study done by Manandhar et al [6] showed Klebsiella spp as most common isolates(34%) followed by Enterobacter (24%).

Rasul et al [18] have pointed out that there is a difference between bacteriological profile in EOS and LOS and also among developing and developed countries. Grampositive organisms constitute a major bulk of etiological agent for both EOS and LOS in developed countries and GBS is seen more in association with EOS. While in developing countries gram negative organisms are responsible for both EOS & LOS, Esch. coli being most commonly associated with EOS. This finding is inconsistent with our findings perhaps, thereby pointing to the fact that this spectrum of flora can have a local variation.

Our study had several limitations, including being a single-centered study with a relatively short duration and a smaller sample size compared to other studies. Additionally, premature babies, low birth weight babies, and most high-risk groups were excluded, which may have influenced the findings. Inter-observer variation in assessments might have also impacted the results, as only purposive sampling was employed. Moving forward, the Modified HSS has the potential to serve as a valuable screening tool for the early diagnosis of neonatal sepsis, but further studies are required to validate its effectiveness and reliability.

## CONCLUSION

Modified HSS is an effective tool for early diagnosis of neonatal sepsis. With all the diagnostic parameter used in combination, modified HSS system has the highest diagnostic accuracy. Nucleated RBC, newly included parameter was more sensitive. Modified HSS has high sensitivity for sepsis when score ≥4 used as a cut off for the presence of sepsis.

Hence, modified HSS can be determined by the easily available hematological parameters from most of the modern analayser. By expanding the parameter base and recalibrating the weightage given for any parameter along with removal of duplications, results in improved specificity of the scores. So, in this study, overall diagnostic ability of Modified HSS is greatly increased.

## Data Availability

All relevant data are within the manuscript and its Supporting Information files.

## LIST OF ABBREVIATIONS

ANC: Absolute Neutrophil Count
APC: Antigen Presenting Cell
APR: Acute Phase Reactant
CBC: Complete Blood Count
CRP: C-Reactive Protein
DAMPs: Damage Associated Molecular Patterns
DC: Dendritic Cells
DLC: Differential Leucocyte Count
EOS: Early Onset Sepsis
GBS: Group B Streptococcus
G-CSF: Granulocyte Colony Stimulating Factor
GM-CSF: Granulocyte Monocyte Colony Stimulating Factor
HSS: Hematological Scoring System
I:T: Immature: Total
ICU: Intensive Care Unit
IFN-γ: Interferon γ
IL: Interleukin
LBP: Lipopolysaccharide binding protein
LOS: Late Onset Sepsis
MBL: Mannan-binding lectin
MSAF: Meconium Stained Amniotic Fluid
NOD: Nucleotide Binding Oligomerization Domain
NPV: Negative Predictive Value
PAMPs: Pathogen Associated Molecular Patterns
PPV: Positive Predictive Value
PMN: Polymorphonuclear
PROM: Premature Rupture of Membrane
PRR: Pathogen Recognition Receptor
RIGI: Retinoic –acid-induible-protein
SAA: Serum Amyloid A
TLC: Total Leucocyte Count
TLRs: Toll like Receptors
TNF-α: Tumour Necrosis Factor α

## Acknowledgements

We sincerely acknowledge the constant support, guidance, and encouragement of our mentors,Dr. Rajan Shah, Dr. Basudha Khanal, and Dr. Lokraj Shah. We are also grateful to the esteemed faculty, seniors, colleagues, and juniors of the Department of Pathology, BPKIHS, for their invaluable contributions. Our heartfelt thanks to Prof. Dr. Surya Raj Niraula and Asst. Prof. Mr. Dharanidhar Baral for their guidance.

